# Parent-reported auditory difficulties in children with ADHD are associated with attentional and fatigue-related vulnerability, not sound avoidance

**DOI:** 10.64898/2026.09.03.26362201

**Authors:** Busra Kose-Ozkan, Omca Guney, Aysegul Gungor Aydin

**Affiliations:** Biomedical Research Institute of New Jersey, Cedar Knolls, NJ, 07927, USA; Basaksehir Cam and Sakura City Hospital, Department of Child and Adolescent Psychiatry, Istanbul, Turkiye

**Keywords:** decreased sound tolerance, hyperacusis, attention deficit hyperactivity disorder

## Abstract

Atypical sensory processing is common in attention-deficit/hyperactivity disorder (ADHD), yet decreased sound tolerance has received little attention, and it remains unclear whether it reflects actual auditory sensitivity or a vulnerability to attentional breakdown under acoustic load. The only prior study provided a single overall estimate, unable to separate sensory symptoms such as avoidance and distress from attentional and fatigue-related difficulties under noisy or demanding conditions. Here, we examined whether parent-reported auditory difficulties in children with ADHD are uniform or concentrated within specific symptom domains, and whether they vary with clinical presentation or medication. A total of 226 children (ages 6-12; 162 with ADHD, 64 typically developing controls) were assessed using the parent-report adaptation of the Khalfa Hyperacusis Questionnaire (Turkish version). The ADHD cohort was stratified by medication status and clinical presentation, and total, subscale, and item-level scores were compared across groups. Our findings show that children with ADHD scored higher than controls on the total and hyperacusis-effects subscale, but not on the coping subscale. Only four of the eleven items distinguished the groups, all reflecting concentration in noise or its decline under fatigue; no item reflecting sound avoidance, sensitivity, or distress differed between groups. This pattern was independent of clinical presentation, medication, and age; it persisted after adjustment for parental education. These results suggest that elevated auditory difficulties in ADHD are better explained by attention- and fatigue-related vulnerability than by heightened auditory sensitivity and may warrant environmental rather than audiological interventions.

## 1. Introduction

Attention-deficit/hyperactivity disorder (ADHD) is one of the most common lifelong neurodevelopmental disorders with a childhood onset. The main characteristics of ADHD involve difficulties with attention and hyperactive or impulsive behaviors, which are classified into three presentations: predominantly inattentive, predominantly hyperactive-impulsive, and combined [1]. Beyond its well-known behavioral and cognitive features, ADHD has been increasingly recognized as a condition that also involves atypical sensory processing: an estimated 50–70% of children with ADHD present with sensory processing difficulties [2–4], compared with 3–16% in general community samples of young children [5,6]. Children with ADHD exhibit more severe sensory atypicalities than controls across multiple domains, including sensitivity, avoidance, low registration, and seeking behaviors [4,7–10]. These atypical sensory experiences are proposed to influence cognitive and psychosocial development [11,12], diminish the quality and enjoyment of social interactions [13], and contribute to difficulties in emotional and attentional regulation [14]. Among various forms of atypical sensory processing, decreased sound tolerance has received limited attention in ADHD research despite its potential to exacerbate attentional and emotional dysregulation.

Hyperacusis is a chronic condition characterized by an abnormally strong reaction to everyday environmental sounds, reflecting reduced tolerance or heightened auditory sensitivity [15]. Critically, hyperacusis is not a unitary construct: it is variably described as auditory hypersensitivity, as well as stress, anxiety, social isolation, and sensory avoidance behaviors triggered by sound [16,17]. This heterogeneity suggests that different components of "sound intolerance" may have distinct correlates and clinical significance. From a neurophysiological perspective, hyperacusis is thought to result from enhanced central gain mechanisms that increase neural responsiveness within the auditory pathways [18]. Through strengthened connections between the auditory cortex and limbic regions, this heightened gain may amplify emotional and attentional reactions to sound, potentially linking auditory hypersensitivity with the hyperactivity, inattention, and impulsivity frequently observed in ADHD [19]. More broadly, sensory over-responsivity, a pattern in which individuals react to sensory input more rapidly, intensely, or for longer duration than their peers [20], is strongly associated with ADHD and Autism Spectrum Disorder [21,22], suggesting that decreased sound tolerance may represent an auditory-specific manifestation of broader sensory regulation difficulties in ADHD.

To date, only one study has specifically examined hyperacusis in ADHD. In a preliminary investigation of 30 children with ADHD and 30 matched controls, Ralli et al. reported a hyperacusis prevalence of 36.7% in the ADHD group compared with 13.3% in controls, using the original Khalfa Hyperacusis Questionnaire [23], a self/clinician-administered instrument developed for adults and subsequently applied to a pediatric sample without a formally validated pediatric parent-report adaptation [19]. This finding provided the first direct evidence linking ADHD to elevated hyperacusis symptoms, but the Khalfa questionnaire yields a single prevalence-based outcome and cannot separate items reflecting sensory avoidance and distress from items reflecting task performance under acoustic or fatigue load. Whether decreased sound tolerance in ADHD reflects heightened sensory sensitivity itself or instead a vulnerability to attentional and fatigue-related breakdown under acoustic load is therefore unknown. These two possibilities carry different mechanistic implications and call for different clinical management. Ralli et al.’s study was also limited to children with the combined presentation, so whether decreased sound tolerance differs across ADHD subtypes remains unknown. It was also limited by a small sample size, and the medication status was neither examined nor stratified to examine the role of pharmacological treatment, a noticeable gap, since stimulants act on catecholaminergic pathways that also modulate arousal and sensory processing [24].

This study had three aims. The first was to determine whether children with ADHD show a greater decrease in sound tolerance than typically developing children, using a validated parent-report instrument capable of distinguishing hyperacusis-related symptoms from avoidance/distress-related symptoms, allowing us to test whether any group differences are uniform across these domains or concentrated in one domain, addressing the ambiguity left unresolved by prior single-outcome instruments. The second was to establish whether decreased sound tolerance differs between the inattentive and combined presentations of ADHD, a question that has not been examined before. The third was to test whether ongoing pharmacological treatment is associated with decreased sound tolerance, a question no previous study has addressed. Clarifying these points has direct clinical relevance. If decreased sound tolerance is elevated in ADHD, specifically in its attention- and fatigue-related components, it may continue to disrupt functioning in classrooms and social settings, even in children whose core symptoms are well controlled by treatment and would warrant a different clinical response than if it reflected genuine sensory hypersensitivity requiring separate audiological assessment.

## 2. Materials and Methods

### 2.1. Study design and ethical approval

The study was approved by the Basaksehir Cam and Sakura City Hospital Ethics Committee (approval no: 2024-KAEK-11, date: 08.04.2025), and it was conducted in accordance with the Declaration of Helsinki and its later amendments. The purpose and scope of the study were explained, and written informed consent was obtained from the parents of all participants prior to enrollment.

### 2.2. Participants

The study included 226 children of both sexes, aged between 6 and 12 years: 162 children with ADHD and 64 typically developing controls comparable in age and sex. ADHD was diagnosed by a child and adolescent psychiatrist according to the criteria of the fifth edition of the Diagnostic and Statistical Manual of Mental Disorders (DSM-5). Within the ADHD cohort, children were characterized along two dimensions. The first was pharmacological treatment, with 81 children receiving medication and 81 receiving none. The second was clinical presentation: 50 children had a predominantly inattentive presentation, and 112 had a combined presentation. Crossing these dimensions yielded four subgroups (unmedicated inattentive, n = 31; unmedicated combined, n = 50; medicated inattentive, n = 19; medicated combined, n = 62). Among the 81 medicated children, 75 (92.6%) were receiving methylphenidate, 11 (13.6%) were receiving atomoxetine, and 5 were receiving both medications. Accordingly, the primary analysis contrasted three groups (unmedicated ADHD, medicated ADHD, and controls), while the effects of clinical presentation and its interaction with medication were examined within the ADHD cohort using the full two-by-two structure.

Hearing status was screened using the parental questionnaire of the national School-Age Hearing Screening Program of the Turkish Ministry of Health, which covers family and medical history relevant to hearing loss (including ototoxic exposure, head trauma, and hereditary conditions) as well as behavioral signs of auditory processing disorder. Exclusion criteria for all participants were a known hearing loss or any otorhinolaryngologic condition that could affect hearing or sound perception, based on parent-reported history, as no audiometric testing was performed as part of this study, a comorbid diagnosis of autism spectrum disorder, developmental delay, intellectual disability, anxiety, depression, speech disorder, or any other neurodevelopmental or psychiatric condition, and incomplete or inconsistent completion of the questionnaires by the parent. For the control group, additional exclusion criteria were a diagnosis of ADHD, use of any neurological or psychiatric medication, and the presence of any chronic illness.

### 2.3. Assessment of decreased sound tolerance

Decreased sound tolerance was assessed using the Turkish version of the Pediatric Hyperacusis Questionnaire, Parent Form (P-HQ). The original 11-item instrument was developed by Carson et al. as a parent-report adaptation of the Khalfa Hyperacusis Questionnaire [25], and its Turkish validity and reliability were established by Öztürk Özdeş et al. [26], who reported high internal consistency (Cronbach’s α = 0.82) and a two-factor structure. Each item is rated on a three-point scale (Yes = 2, Sometimes = 1, No = 0), with no reverse-scored items, yielding a total score from 0 to 22; higher scores indicate decreased sound tolerance. The scale comprises two subscales, as established in the Turkish validation study: Factor 1, reflecting the effects of hyperacusis (items 1, 2, 4, 5, 10, and 11), and Factor 2, reflecting coping and social relations (items 3, 6, 7, 8, and 9). Throughout this manuscript, we use decreased sound tolerance to refer to P-HQ scores as measured, the operational construct captured by parent report on this instrument, and reserve “sound intolerance” for our interpretation of what these scores reflect, as developed in the Discussion.

For children with ADHD, parents completed a demographic information form and the P-HQ in paper format during a clinic visit at the Child and Adolescent Psychiatry outpatient clinic of the Basaksehir Cam and Sakura City Hospital. The control group was recruited from the community using an online version of the same forms; an initial group of families was invited to participate and asked to share the survey link with other families in their social networks who had typically developing children of comparable age, following a snowball sampling approach [27]. In both groups, a single parent completed one questionnaire per child, and only fully completed forms were included in the analysis. For the medicated ADHD group, parents rated their child’s behavior during ongoing pharmacological treatment.

### 2.4. Statistical analyses

Analyses were performed in IBM SPSS Statistics version 28 (IBM Corp., Armonk, NY, USA) and graphs in GraphPad Prism version 11 (GraphPad Software, San Diego, CA, USA). Continuous variables were summarized as mean ± standard deviation, and categorical variables as frequencies. Normality was assessed within each group using the Shapiro-Wilk test for every outcome variable. Because the normality assumption was violated in at least one group for every outcome, non-parametric tests were used for the primary between-group comparisons; corresponding parametric analyses (independent t-tests/one-way ANOVA in place of Mann-Whitney U/Kruskal-Wallis) were run in parallel as a robustness check and are reported in Supplementary Table S1 where results diverged from the non-parametric findings. A two-sided p < .05 was considered significant.

Group comparability in age and sex was assessed using the Kruskal-Wallis and chi-square tests. For categorical comparisons involving cells with expected counts below 5 (e.g., family psychiatric history in the control group), Fisher’s exact test was used in place of chi-square.

The primary analysis compared P-HQ total and subscale (Factors 1 and 2) scores across the three groups (unmedicated ADHD, medicated ADHD, control) using the Kruskal-Wallis test with Bonferroni-adjusted pairwise comparisons and effect sizes expressed as Cohen’s d. To identify the source of the group differences, each of the 11 P-HQ items was compared across groups using the Kruskal-Wallis test with Bonferroni correction applied across the 11 items (adjusted α = .0045).

Four additional analyses were conducted to test the robustness of the primary findings and to address the study’s third aim (1) a chi-square comparison of the proportion of children scoring above the previously reported clinical cut-off of 10 points [25] across the three groups, (2) Spearman correlations between age and P-HQ total scores within each group, with Bonferroni correction applied across the three groups (adjusted α = .017); (3) an analysis of covariance (ANCOVA) testing the primary group effect on P-HQ total and Factor 1 scores with maternal and paternal education entered as covariates, to address the observed group difference in parental education; and (4) a two-way ANOVA (medication × clinical presentation) on P-HQ total and subscale scores within the ADHD cohort, corresponding to the study’s third aim of testing whether pharmacological treatment is associated with decreased sound tolerance. Where results from these four analyses are interpreted jointly against a stricter threshold, a Bonferroni-corrected α of .0125 (.05/4) is reported alongside the nominal p-value.

Effect sizes are reported throughout as Cohen’s d for pairwise non-parametric comparisons and partial η² for ANOVA and ANCOVA models, to allow consistent comparison of effect magnitude across the primary and additional analyses. A sensitivity power analysis (G*Power 3.1.9.4) indicated that, with the available ADHD sample (n = 162), the two-way analysis had 80% power (α = .05) to detect main effects and an interaction of Cohen’s f ≥ 0.22 (partial η² ≥ .047), corresponding to a small-to-medium effect. This level of power applies to the main effects, which draw on the full sample. The interaction, however, is estimated less efficiently because of the unequal cell sizes (the smallest being the medicated-inattentive subgroup, n = 19); accounting for this imbalance, the achieved power to detect an effect of f = 0.22 was approximately 72%. Non-significant interaction effects should therefore be interpreted as inconclusive rather than as confirmatory evidence of no interaction.

## 3. Results

### 3.1. Descriptive statistics

Table 1 presents the demographic characteristics of the three groups. The groups were comparable in age (p = .438) and sex distribution (p = .319), with boys predominating in all three groups. Maternal and paternal ages did not differ meaningfully across groups, although maternal age differed significantly (p = .028). The groups did, however, differ in parental education (both p < .001, Kruskal-Wallis test). University-level education or higher was substantially more common among the parents of control children (mothers 57.8%, fathers 61.0%) than among the parents of children with ADHD (mothers 18.5% in the unmedicated and 23.5% in the medicated group; fathers 14.8% and 22.2%, respectively). Its potential confounding effect on P-HQ scores was therefore examined directly in a subsequent analysis of covariance (**See section 3.5. Additional analyses**). The family history of psychiatric disorder was also more frequent in the ADHD groups than in controls (Fisher’s exact test, p = .017), consistent with the known heritability of ADHD.

**Table 1.** Demographic characteristics of the study participants.

|  | **Unmedicated**<br>**ADHD (n=81)** | **Medicated**<br>**ADHD (n=81)** | **Control**<br>**(n=64)** | **p** |
| --- | --- | --- | --- | --- |
| **Age, years (mean ± SD)** | 8.80 ± 1.83 | 8.52 ± 1.75 | 8.41 ± 1.69 | .438 |
| **Male, n (%)** | 64 (79.0) | 65 (80.2) | 45 (70.3) | .319 |
| **Maternal age, years (mean ± SD)** | 35.49 ± 5.25 | 37.31 ± 6.11 | 37.97 ± 5.62 | .028 |
| **Paternal age, years (mean ± SD)** | 39.05 ± 5.88 | 40.47 ± 6.92 | 40.86 ± 5.98 | .148 |
| **Family psychiatric history, n (%)** | 8 (9.9) | 12 (14.8) | 1 (1.6) | .017 |
| **Maternal education, n (%)** |  |  |  | **<.001** |
| Primary | 20 (24.7) | 16 (19.8) | 6 (9.4) |  |
| Secondary | 19 (23.5) | 16 (19.8) | 1 (1.6) |  |
| High school | 26 (32.1) | 28 (34.6) | 20 (31.2) |  |
| University | 14 (17.3) | 17 (21.0) | 31 (48.4) |  |
| Postgraduate | 1 (1.2) | 2 (2.5) | 6 (9.4) |  |
| **Paternal education, n (%)** |  |  |  | **<.001** |
| Primary | 18 (22.2) | 15 (18.5) | 3 (4.7) |  |
| Secondary | 26 (32.1) | 21 (25.9) | 6 (9.4) |  |
| High school | 25 (30.9) | 27 (33.3) | 16 (25.0) |  |
| University | 10 (12.3) | 15 (18.5) | 30 (46.9) |  |
| Postgraduate | 2 (2.5) | 3 (3.7) | 9 (14.1) |  |

### 3.2. Distribution of P-HQ scores

Normality was assessed with the Shapiro-Wilk test within each group. For the P-HQ total score, the distribution departed from normality in the medicated ADHD group (W = .962, p = .017) and in the control group (W = .916, p < .001), but not in the unmedicated ADHD group (W = .977, p = .154). For Factor 1, only the control group deviated from normality (W = .903, p < .001; unmedicated ADHD W = .974, p = .092; medicated ADHD W = .975, p = .112). Factor 2 scores were non-normally distributed in all three groups (W = .820, .808, and .774, respectively; all p < .001), reflecting a pronounced positive skew. Because the normality assumption was violated in at least one group for every outcome variable, non-parametric tests were used for the primary between-group comparisons. Parametric analyses were additionally performed and yielded the same pattern of results.

### 3.3. Decreased sound tolerance across groups

P-HQ total scores differed significantly across the three groups (Kruskal-Wallis H = 31.44, df = 2, p < .001). Both ADHD groups scored higher than controls: unmedicated ADHD 8.05 ± 3.93 and medicated ADHD 7.77 ± 4.48, compared with 4.39 ± 3.76 in controls. Pairwise comparisons with Bonferroni correction confirmed that both the unmedicated (adjusted p < .001, d = 0.95) and the medicated ADHD group (adjusted p < .001, d = 0.81) differed from controls, whereas the two ADHD groups did not differ from each other (adjusted p = 1.000, d = 0.07).

The same pattern was observed for Factor 1 (effects of hyperacusis), where group differences were even more pronounced (H = 40.52, df = 2, p < .001; unmedicated 6.32 ± 2.89, medicated 6.05 ± 2.95, controls 3.25 ± 2.83). Both ADHD groups again exceeded controls (adjusted p < .001 for both), with no difference between them (adjusted p = 1.000). In contrast, no group differences were found for Factor 2 (coping and social relations; H = 2.39, df = 2, p = .302; unmedicated 1.73 ± 2.03, medicated 1.72 ± 2.08, controls 1.14 ± 1.41).

The three groups were comparable in sex distribution (χ² = 2.28, df = 2, p = .319), with boys predominating in all groups (79.0%, 80.2%, and 70.3% in the unmedicated ADHD, medicated ADHD, and control groups, respectively). P-HQ total scores did not differ between girls and boys (6.54 ± 4.18 vs 7.02 ± 4.43; U = 4794.5, p = .512), and no sex difference was found for either subscale (Factor 1: U = 4540.0, p = .969; Factor 2: U = 5146.5, p = .115). A two-way analysis of variance confirmed a significant main effect of group (F(2, 220) = 15.99, p < .001, partial η² = .127) but no main effect of sex (F(1, 220) = 0.02, p = .891) and no group × sex interaction (F(2, 220) = 1.87, p = .157), indicating that group differences in sound tolerance were independent of sex.

### 3.4. Item-level analysis

To identify which aspects of the questionnaire drove the group differences, each of the 11 P-HQ items was compared across the three groups, grouped by subscale. After Bonferroni correction across the 11 items, all four significant items belonged to Factor 1 (Figure 1). Two reflected functioning specifically in noisy conditions: concentrating in noise (item 1, H = 38.54, p < .001) and reading in noise (item 2, H = 37.73, p < .001). The other two reflected the susceptibility of concentration to fatigue: a decline in concentration toward the end of the day (item 10, H = 27.27, p < .001) and the impact of stress and fatigue on concentration (item 11, H = 19.03, p < .001). On all four items, both ADHD groups scored higher than controls (p < .05), whereas the medicated and unmedicated groups never differed from each other. Notably, the two remaining Factor 1 items did not distinguish the groups: difficulty ignoring everyday sounds (item 4) and sensitivity to street noise (item 5) were comparable across groups.

**Figure 1.**
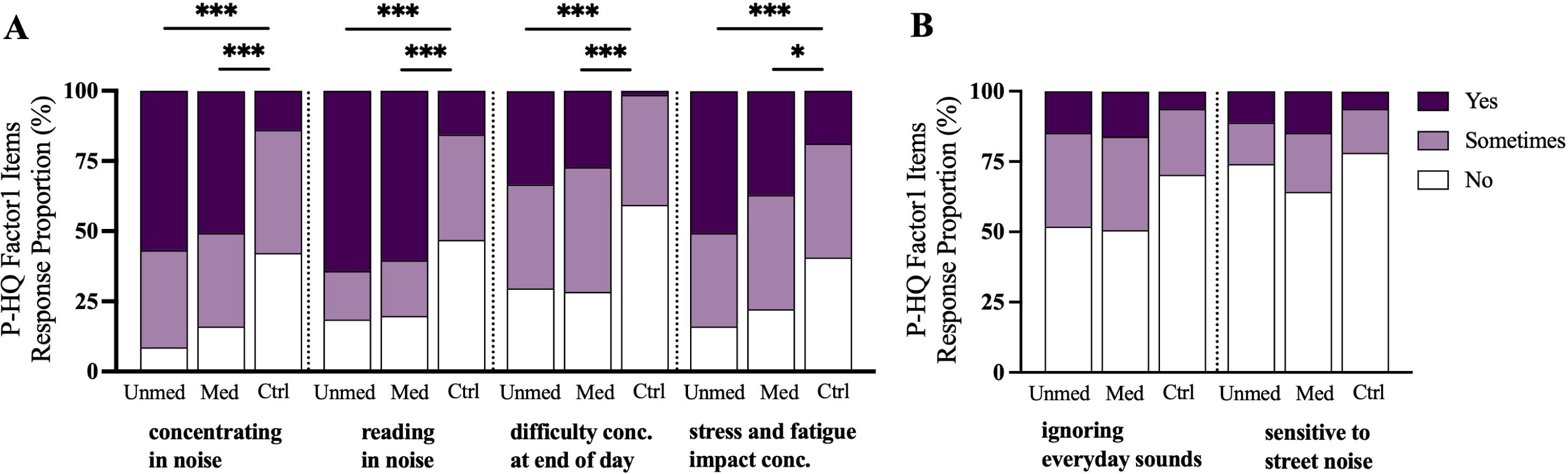
Item-level P-HQ scores for Factor 1 (effects of hyperacusis) items across the three groups. Stacked bars show the proportion of parents who responded "Yes," "Sometimes," or "No" to each item, for the unmedicated ADHD, medicated ADHD, and control groups. Group differences were tested with the Kruskal-Wallis test, with Bonferroni correction across the 11 items. (A) The four items that differed significantly between groups were: concentrating in noise, reading in noise, difficulty concentrating at the end of the day, and stress and fatigue impact on concentration. On all four, both ADHD groups reported more frequent difficulties than controls, while the two ADHD groups did not differ. (B) The two remaining Factor 1 items (ignoring everyday sounds; sensitive to street noise) did not differ across groups. Asterisks indicate significant pairwise differences from controls (Bonferroni-corrected): *p < .05, ***p < .001.

None of the five Factor 2 items (coping and social relations) differed across groups (Figure 2), including the use of ear protection (item 3), turning down invitations because of noise (item 6), being bothered by noise in social situations (item 7), poor tolerance noticed by others (item 8), and sound-induced stress or irritation (item 9; all p = 1.000 except item 8, p = .649). Thus, across the entire questionnaire, group differences were restricted to items reflecting attention and concentration and were absent from every item reflecting heightened sensitivity to, avoidance of, or distress from sound.

**Figure 2.**
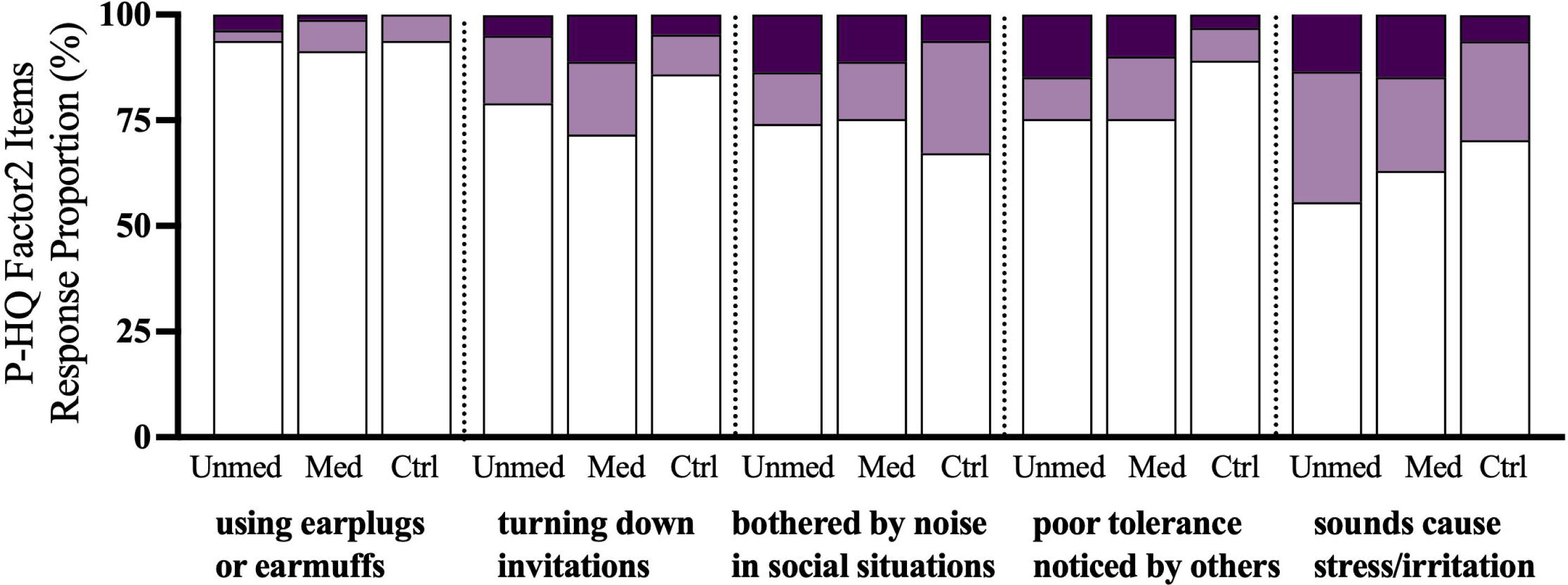
Item-level P-HQ scores for Factor 2 (coping and social relations) items across the three groups. Stacked bars show the proportion of parents who responded "Yes," "Sometimes," or "No" to each item, for the unmedicated ADHD, medicated ADHD, and control groups. The five items shown are: using earplugs or earmuffs, turning down invitations because of noise, being bothered by noise in social situations, poor tolerance noticed by others, and sounds causing stress or irritation. None of the five items differed significantly across groups (Kruskal-Wallis test with Bonferroni correction across the 11 items; all p > .05).

### 3.5. Additional analyses

#### Prevalence based on the P-HQ cut-off

As a secondary analysis, children were classified using the previously reported cut-off of 10 points. More children in the unmedicated (21.0%) and medicated ADHD groups (24.7%) scored above the threshold than in controls (7.8%), a difference that was significant across the three groups (χ² = 7.21, df = 2, p = .027). Since the two ADHD groups did not differ, they were combined to compare ADHD with controls: 22.8% of children with ADHD scored above the cut-off, compared with 7.8% of controls (χ² = 5.89, p = .015).

#### Association with age

The relationship between age and P-HQ total score was examined separately within each group (Figure 3). Scores were unrelated to age in the unmedicated ADHD group (rho = .02, uncorrected p = .858, corrected p = 1.000) and showed a weak negative trend in the control group (rho = -.11, uncorrected p = .397, corrected p = 1.000). In the medicated ADHD group, a positive association was observed (rho = .24, uncorrected p = .031), but this did not survive Bonferroni correction for the three comparisons (corrected p = .093). Decreased sound tolerance, therefore, showed no statistically reliable association with age in any group.

**Figure 3.**
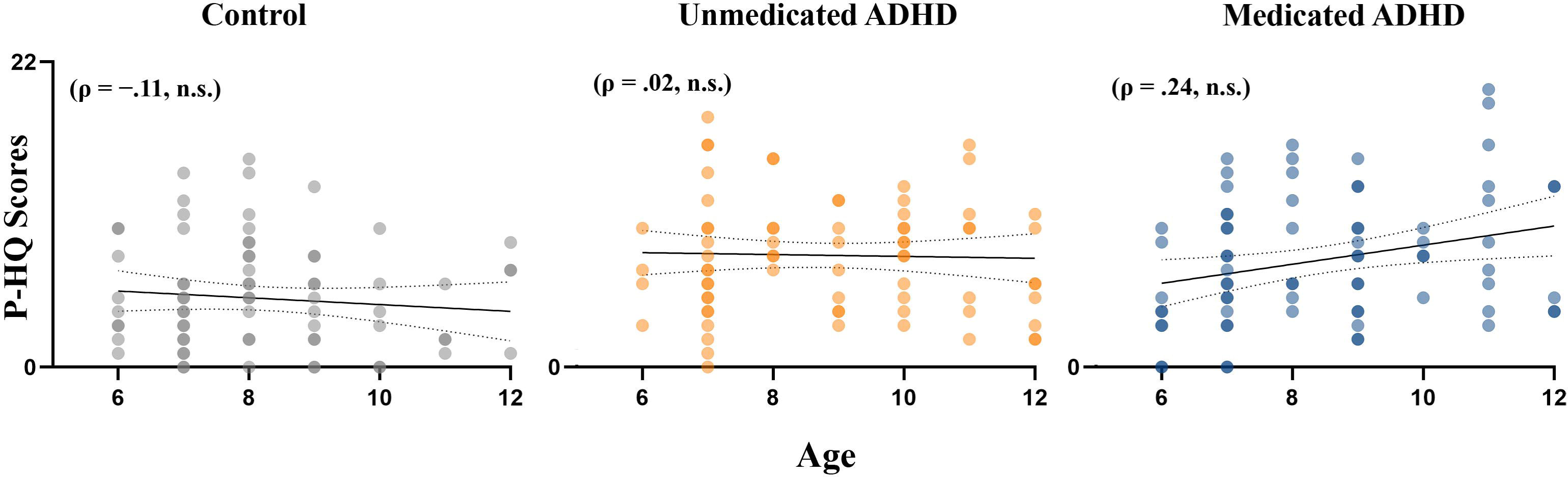
Age and P-HQ total scores in the three groups. Each panel shows individual scores for the control, unmedicated ADHD, and medicated ADHD groups, with a linear regression line and 95% confidence band (dotted lines). Point opacity was reduced to indicate overlapping data points, with darker points reflecting a greater number of children sharing the same age and score. Spearman correlations were non-significant in all groups after Bonferroni correction (control rho = -.11, p = 1.000; unmedicated ADHD rho = .02, p = 1.000; medicated ADHD rho = .24, p = .093).

#### Effect of parental education

Because parental education differed between the clinically recruited ADHD groups and the community-recruited controls, its potential confounding effect was assessed using analysis of covariance with maternal and paternal education as covariates. For the P-HQ total score, the group effect remained significant after adjustment (F(2, 221) = 15.25, p < .001, partial η² = .121), and neither maternal (F(1, 221) = 2.37, p = .125, partial η² = .011) nor paternal education (F(1, 221) = 0.66, p = .416, partial η² = .003) predicted scores. The same held for Factor 1 (group effect F(2, 221) = 21.77, p < .001, partial η² = .165). Adjusted marginal means closely matched the unadjusted values, and both ADHD groups continued to exceed controls (p < .001) with no difference between them (p = 1.000). Group differences in decreased sound tolerance were therefore independent of parental education.

#### Effect of ADHD presentation and medication

Within the ADHD cohort, a two-way analysis (medication × presentation) revealed no effect of clinical presentation on P-HQ scores. Inattentive and combined presentations did not differ on the total score (8.18 ± 4.21 vs 7.79 ± 4.22; p = .757), Factor 1 (p = .556), or Factor 2 (p = .538), and the presentation main effect remained non-significant when medication was controlled (total: F(1, 158) = 0.38, p = .536, partial η² = .002; Factor 1: F(1, 158) = 1.01, p = .317, partial η² = .006; Factor 2: F(1, 158) = 0.02, p = .884, partial η² < .001). Medication status showed no main effect (total: F(1, 158) = 0.01, p = .926, partial η² < .001), and no medication × presentation interaction reached significance (total F(1, 158) = 1.03, p = .313, partial η² = .006; Factor 1:F(1, 158) = 2.97, p = .087, partial η² = .018; Factor 2:F(1, 158) = 0.12, p = .725, partial η² = .001). Because the sample was large enough to detect at least small-to-medium effects (f ≥ 0.22, partial η² ≥ .047), and the observed effects were substantially smaller than this (partial η² ≤ .018), the absence of significant differences is unlikely to reflect insufficient statistical power. Decreased sound tolerance in ADHD was therefore independent of both clinical presentation and pharmacological treatment. When a Bonferroni correction is applied jointly across the four additional analyses (α = .0125), the prevalence-cutoff difference between ADHD and control groups no longer reaches significance and should be interpreted as suggestive rather than confirmatory. The ANCOVA-adjusted group effect on P-HQ total score remains significant at this stricter threshold (p < .001).

## 4. Discussion

The central finding of this study is not that children with ADHD show a generalized decrease in sound tolerance, but that the elevated scores driving this pattern are concentrated entirely on items reflecting attention and fatigue-related vulnerability, not on sound sensitivity or avoidance itself. Four individual P-HQ items, difficulty concentrating in noise, difficulty reading in noise, decline in concentration by day’s end, and susceptibility of concentration to stress and fatigue, that differed significantly between ADHD and control groups after correction, all belong to Factor 1 and describe cognitive performance degrading under acoustic or fatigue load. In contrast, every Factor 2 item, capturing the behavioral signature of true sensory over-responsivity, sound avoidance, use of ear protection, social withdrawal due to noise, and sound-induced distress, showed no group difference. Even within Factor 1, two items describing sensitivity to sound itself rather than its effect on concentration (difficulty ignoring everyday sounds, sensitivity to street noise) failed to distinguish the groups.

This dissociation is difficult to reconcile with a straightforward interpretation of hyperacusis. True auditory hypersensitivity, an abnormally strong physiological or affective reaction to sound [17,16], would be expected to manifest in avoidance, distress, and coping behavior, which is what Factor 2 measures. Its absence, combined with the highly specific elevated scores on attention- and fatigue-linked items, points toward a different mechanism. Rather than reduced tolerance for sound itself, children with ADHD may have attentional resources that are more easily depleted by competing auditory input, particularly under fatigue. We note that this dissociation could equally reflect selective informant bias rather than a true absence of sensory avoidance behavior. Parents of children with ADHD may be primed to notice and report attention-related difficulties given the nature of the diagnosis itself, independent of the child’s underlying sensory experience. Two findings from the study push this interpretation beyond a descriptive item-level pattern and toward a specific mechanistic claim, though both should be read as suggestive rather than conclusive. Medication status showed no association with P-HQ total and subscale (Factors 1 and 2) scores, and no interaction with clinical presentation. Stimulants act directly on sustained attention [24,28], so if this vulnerability were simply a downstream symptom of unmedicated inattention, some attenuation with treatment might be expected. Its absence is consistent with a more stable, trait like feature of ADHD that persists regardless of treatment status, though a null result here is equally consistent with partial and variable treatment response, incomplete symptom control despite medication, or a parent-report measure insensitive to treatment-related change within this sample. What that stable feature consists of, however, is a separate question. The P-HQ does not measure hyperacusis in the clinical sense. Clinical hyperacusis denotes a measurable reduction in loudness tolerance, confirmed through objective testing such as loudness discomfort level (LDL) assessment [29], and is attributed to altered central auditory gain [18]. The P-HQ captures parent-perceived difficulty with sound-related situations, a construct better described as subjective sound intolerance, which may or may not correspond to an underlying change in auditory perception. Without audiometric or LDL testing in the present study, we cannot determine whether elevated P-HQ scores reflect a true reduction in loudness tolerance or a purely perceptual and attentional phenomenon that leaves auditory processing itself unaffected. Given that our item-level, medication, and presentation findings converge on the latter interpretation, but rely solely on parent-report data, we use the term "sound intolerance" rather than "hyperacusis" throughout this discussion to reflect the level of evidence our study actually supports. This reframes the prior literature’s characterization of "hyperacusis in ADHD" [19] as potentially an attentional rather than a sensory phenomenon. Ralli et al. included an audiological evaluation to confirm normal hearing, not to directly quantify loudness tolerance. Their hyperacusis diagnosis required positivity on both the parent questionnaire and the child interview, a dual-informant subjective criterion, not an objective one, and relied on the original Khalfa Hyperacusis Questionnaire rather than the pediatric parent-report P-HQ used in our study, and the two may not capture identical constructs. Most importantly, their single binary outcome could not have detected the domain-level dissociation reported here, even if the same pattern were present in their sample.

This pattern is consistent with the broader literature on sensory processing in ADHD, which reports atypicalities that are elevated overall but heterogeneous across domains rather than uniform impairment [4,30,31]. Meta-analytic and comparative work has found that children with ADHD show elevated sensory sensitivity, avoidance, and low registration compared to neurotypical peers across multiple modalities [4,7], yet the specific domains and item types affected vary considerably across studies and sensory channels, including tactile processing [32] and cross-modal over-responsivity linked to attentional capture [33]. Sensory over-responsivity itself has been conceptualized less as a unified sensory deficit than as a regulatory difficulty in filtering or modulating input under cognitive or emotional load [20], a framing consistent with the present findings, in which auditory complaints emerged specifically under conditions of competing demand (noise during a concentration task) or fatigue (decline by day’s end), rather than as a generalized aversion to sound. This interpretation aligns with frameworks of effortful listening, which hold that understanding speech under adverse conditions, such as background noise, draws on limited cognitive resources beyond those required in quiet [34]. Consistent with this, children with ADHD have been shown to experience greater difficulty than typically developing peers on speech-in-noise tasks, a difficulty linked to their reduced working memory capacity [35]. Because parsing speech from competing noise is attentionally demanding, children with ADHD, whose attentional and working-memory resources are already constrained, may experience disproportionate difficulty and fatigue in noisy environments even without elevated auditory sensitivity. This would account for the specific pattern observed here, in which the items distinguishing children with ADHD concerned concentration in noise and its decline under fatigue, rather than sensitivity to sound itself. Taken together with prior reports linking sensory modulation difficulties to attentional and behavioral dysregulation in ADHD [21,22], our findings suggest that auditory difficulties in this population are better understood as one manifestation of a broader regulatory vulnerability under load that persists independent of ADHD pharmacological treatment, rather than as a domain-specific sensory abnormality analogous to hyperacusis in its clinical sense.

These two possible explanations, a domain-specific sensory abnormality versus a broader regulatory vulnerability expressed under load, carry direct and divergent clinical implications. If sound intolerance in ADHD reflected true auditory hypersensitivity, the appropriate response would be audiological referral and evaluation for hyperacusis-specific interventions such as sound therapy or desensitization protocols. If, instead, it reflects attentional resources being more easily depleted by competing acoustic input under fatigue, the pattern our item-level, medication, and presentation findings collectively support, the more relevant response shifts toward environmental and behavioral accommodations aimed at supporting attention rather than treating a sensory disorder: reducing background noise in classroom or homework settings, scheduling demanding cognitive tasks earlier in the day before fatigue-related decline sets in, and building in structured breaks during noisy or high-demand periods. Given the absence of audiometric confirmation in this study, clinicians should not rule out hyperacusis on this basis alone, particularly in children with persistent or severe sound-related distress, for whom an audiological evaluation remains warranted. But for the majority of children with ADHD presenting with sound-related complaints in the absence of avoidance or distress, our findings suggest the regulatory pathway, not the sensory one, is the more likely explanation, and that this vulnerability should not be assumed to resolve with pharmacological treatment of core ADHD symptoms alone.

## Limitations

Several limitations of the present study need to be considered when interpreting the findings. First, we did not use audiological methods such as LDL testing or acoustic reflex measures to assess decreased sound tolerance. Their applicability to the pediatric population remains debated, given reliability concerns [36,37] and inconsistent testing protocols across studies [38,39]. Instead, decreased sound tolerance was assessed using a parent-report questionnaire, a Turkish adaptation of the Khalfa Hyperacusis Questionnaire [23], validated by Öztürk Özdeş et al. [26]. This reliance on parent-report measures also introduces the risk of informant bias, as parents’ ratings may be shaped by their awareness of their child’s diagnostic status. Second, the control group was recruited from the community through snowball sampling, whereas the ADHD groups were recruited from a clinical setting. We addressed this limitation with additional analyses. Third, because only children with a diagnosis of ADHD without co-occurring conditions were included in our study samples, our findings should not be extrapolated to children with co-occurring conditions. Fourth, Full-Scale Intelligence Quotient was not assessed and could not be examined as a covariate, so we cannot rule out the possibility that unmeasured differences in cognitive ability contributed to the attentional pattern observed here. Addressing these limitations, through objective audiometric assessment, multi-informant designs incorporating raters blind to diagnostic status, such as teacher report, inclusion of children with co-occurring conditions, and assessment of cognitive ability, represent the necessary next step for determining whether the attentional dissociation observed here reflects a stable and generalizable feature of ADHD-related sound intolerance.

## Data Availability

All data produced in the present study are available upon reasonable request to the authors

## Author contributions

Conceptualization: B.K.O. and O.G.; Methodology: B.K.O., O.G., A.G.A.; Investigation: O.G.; Formal analysis: B.K.O. and A.G.A.; Writing –original draft: B.K.O. and A.G.A.; Writing –review and editing: B.K.O. and A.G.A. All authors read and approved the manuscript.

