## Supplemental Table 1 for "Parent-reported auditory difficulties in children with ADHD are associated with attentional and fatigue-related vulnerability, not sound avoidance"

### Supplementary Table 1

*Concordance between non-parametric and parametric analyses.*

Panel A. Primary three-group comparison

| Outcome | H (KW) | p | ε² | F (ANOVA) | p | η² |
| --- | --- | --- | --- | --- | --- | --- |
| P-HQ total | 31.44 | <.001 | 0.132 | 17.04 | <.001 | 0.133 |
| Factor 1 | 40.52 | <.001 | 0.173 | 23.70 | <.001 | 0.175 |
| Factor 2 | 2.39 | 0.302 | 0.002 | 2.16 | 0.118 | 0.019 |

Panel B. Two-way ANOVA main effects (within ADHD cohort)

| Outcome (effect) | U (MWU) | p | F (ANOVA) | p | η² |
| --- | --- | --- | --- | --- | --- |
| P-HQ total (Medication) | 3484 | 0.494 | 0.01 | 0.926 | 0.000 |
| P-HQ total (Presentation) | 2886 | 0.757 | 0.38 | 0.536 | 0.002 |
| Factor 1 (Medication) | 3444 | 0.582 | 0.07 | 0.787 | 0.000 |
| Factor 1 (Presentation) | 2962 | 0.556 | 1.01 | 0.317 | 0.006 |
| Factor 2 (Medication) | 3315 | 0.906 | 0.04 | 0.849 | 0.000 |
| Factor 2 (Presentation) | 2964 | 0.538 | 0.02 | 0.884 | 0.000 |

KW, Kruskal-Wallis; MWU, Mann-Whitney U; ε², epsilon-squared; η², eta-squared. Parametric and non-parametric tests yielded concordant conclusions for every comparison.
